# Feasibility study of gait analysis using a new Wearable Force Plate

**DOI:** 10.64898/2026.08.30.26361786

**Authors:** Clara B. Sanz-Morère, Gonzalo Garrido-López, Mizuka Hayase, Javier Rueda, Qi An, Shingo Shimoda, Juan C. Moreno, Enrique Navarro

## Abstract

Static force plates (FP) are the gold standard for measuring ground reaction forces (GRF) and computing joint moments through inverse dynamics in gait analysis. However, they are restricted to controlled environments, and the number of steps analyzed is limited by the plates embedded in the floor. To address these limitations, portable solutions such as sensorized insoles, socks, or shoes have emerged. Yet, creating wearable systems capable of measuring three-dimensional GRF in real-world conditions remains challenging. Current sensorized shoes often incorporate thick sensors (up to 2 cm), reducing usability and limiting their application in pathological populations or dynamic tasks like running.

This study evaluates the usability of ShokacShoes, a novel sensorized shoe integrating three thin, three-dimensional force sensors, and explores its potential as a Wearable Force Plate (WFP). Eight healthy participants performed slow, natural, and fast walking using two insole configurations. Force and temporal metrics were derived from WFP and FP data. Results indicate that WFP enables accurate step segmentation and detects significant effects of speed and insole type on temporal and force metrics, confirming its reliability under different walking conditions.

Comparisons with FP revealed differences in force metrics and signal morphology, though temporal parameters remained consistent. These results are likely due to sensor quantity and positioning.

Thereby, ShokacShoes represent a valid solution capable of measuring three-dimensional forces within commercial footwear. Future work will focus on validating the applicability of a new version of ShokacShoes against gold-standard FP in a comprehensive validation study involving diverse real-world scenarios and pathological conditions.

## Introduction

The study of human forces started at the end of the 19th century, with the design of a “research shoe” based on air chambers that was able to reveal the vertical force pattern and its amplitude during walking ^1^. Though this idea held great potential, the measurement of forces during walking was proven easier when using a static force plate (FP) integrated inside the walking floor ^2^ and embedding several sensors with 6 degrees-of-freedom able to measure three-dimensional forces and moments. Ground reaction forces (GRF) measured by means of FP have been proven pivotal to assist in optimizing traumatic rehabilitation outcomes such as after anterior cruciate ligament reconstruction ^3^, to correlate with walking and running speed ^4^, or to diagnose and assess early-stage Parkinson’s disease ^5^. They have also been used to retrieve the Center of Pressure, one of the most common variables used to assess balance control in sports and neurological applications ^6,7^.

Moreover, in the last years, thanks to the evolution of computer power, kinematics analysis combined with GRF and inertial parameters, have been used to compute joint moments using inverse dynamics, which has allowed, among others, to predict injury and fall risks ^8,9^. Currently, static FP are considered the gold standard for accurately assessing gait dynamics, which is crucial to avoid incorrect clinical conclusions ^10,11^. However, their application is generally confined to controlled environments, which allows to isolate the measurements from confounding factors but prevents the evaluation of gait biomechanics in everyday-life and thus limits the generalizability of the results. This constraint also applies to gold-standard optoelectronic systems ^11^. Furthermore, when analyzing GRF, the number of gait cycles that can be assessed within a full corridor is restricted by the number of force plates embedded in the lab floor, which usually ranges from one to four.

As a result, gait analysis sessions must be extended to capture a sufficient number of steps, increasing the duration of the assessment and contributing to patient exhaustion ^12^. To overcome the limitations of current static FP, some solutions have been designed. This reflects a general trend also seen in the emergence of inertial measurement units and markerless motion capture systems, which were introduced to overcome the constraints of traditional optoelectronic technologies.

To replace FP, some solutions are based on pressure mats, such as GAITRite (CIR Systems, New Jersey, USA), which allows the recording of the vertical pressure exerted by the subjects during several steps within the same corridor, but do not provide measurements of three-dimensional forces ^13^. Another alternative is instrumented treadmills, such as the GaitWay3D (h/p/cosmos sports &medical gmbh, Nussdorf-Traunstein, Germany). However, these systems restrict gait analysis to controlled laboratory environments, require considerable space, and limit assessments to treadmill walking, which differs from overground walking ^14^. Additionally, treadmill-based gait assessment can be challenging for individuals with severe gait impairments, particularly those with limited balance or who require external assistance. Other researchers reacquired the idea of the “research shoe” and designed pressure insoles that can be integrated inside each individual’s shoe to record the pressure during walking. Now several of these are commercialized: OpenGo Sensor Insoles (Moticon ReGo AG, Munich, Germany), Pedar (novel Gmbh, Munich, Germany), F-Scan Go (Tekscan, Norwood, MA, United States), Medilogic WLAN insole (Medilogic Gmbh, Schönefeld, Germnay) or Intelligent Insoles | Clinical (XSENSOR® Technology Corporation, Calgary, Canada). This technology allows the estimation of vertical GRF and its analysis with a portable setup but presents very wide technical characteristics: a variable number of pressure sensors ranging from 16 (OpenGo) to 240 (Medilogic); a maximum vertical load ranging from 400 kPa (ReGo) to 883 kPa (Intelligent Insoles |Clinical); a sampling frequency of around 110 Hz, reaching 400 Hz in the sports version of Medilogic WLANInsole. Some insoles are not computer-dependent, with an autonomy of several hours (Intelligent Insoles | Clinical), whereas others rely on a computer or a tablet (Medilogic WLAN insole). Similar technologies have been implemented in smart garments, such as smart socks such as DAid® Pressure Sock System ^15^ and Sensoria Smart socks (Sensoria Fitness, Redmond, WA, United States), aimed at increasing comfort, allowing biomechanical analysis without shoes, and ensuring a natural walking pattern ^15^.

However, relying exclusively on vertical GRF estimation may be insufficient to fully characterize an individual’s biomechanical requirements. Measuring the three-dimensional components of the GRF (vertical force and propulsive and medio-lateral shear forces) is crucial for diagnosing pathological gait ^16,17^, assessing balance ^17^, preventing injuries in athletes and neurologically-compromised individuals ^18,19^, and refining therapeutic interventions ^20^. Moreover, three-dimensional forces are necessary to retrieve joint moments by means of inverse dynamics. Yet, developing portable and usable sensorized shoes that allow measuring three-dimensional GRF and moments in uncontrolled scenarios remains a challenge.

Several attempts have been made in the last years. The first attempt was made by Veltink et al. in 2005 ^12^, who designed the ForceShoes, which included two three-dimensional force and torque sensors under the heel and forefoot and a miniature inertial measurement unit. The shoes were validated against gold-standard FP and tested with stroke survivors ^21,22^. The M3D shoes, designed by Liu et al., in 2010 ^23^, included six three-dimensional force sensors under the heel and forefoot, two three-dimensional accelerometers, and six 1D gyroscopes. The shoes were validated against a FP ^24^. Similar attempts have been made in the last years ^25–27^. Though these validation results were optimistic, the thickness of the sensors ranged from 9.2 to 21 mm, limiting the usability of the shoes and impeding their use in pathological population or in more complex tasks such as running or fast walking. In 2023, Snyder et al. ^28^ designed a sensorized insole using a novel sensor named ShokacChip. They included 5 of these sensors: one under the heel, 3 under the forefoot, and one under the toe. This design allowed for a much thinner insole of only 3.4 mm, included in a conventional shoe. However, the usability was limited by the electronics design, which included visible cable connections. The system was used in a pilot study in which nine healthy females walked over a corridor with 10 embedded FP. The data from the sensorized shoes were used to estimate knee adduction moments, which can be considered a risk factor of knee osteoarthritis, based on machine learning models. However, the raw data of the forces and their comparison against gold-standard forces were not provided in the study. In 2022, Nakai et al. introduced the ShokacShoes ^29^, a new sensorized shoe that outperforms the design of existing technologies and paves the way for portable and high-precision analysis of GRF.

The aim of this feasibility study is to evaluate the usability of the ShokacShoes and to provide an initial biomechanical analysis of gait, highlighting their potential to function as a Wearable Force Plate (WFP). Specifically, we examine the ability of the ShokacShoes to capture three-dimensional GRF during walking in healthy individuals, using three independent sensors across three walking speeds and two insole configurations. We then provide an initial comparison of the forces recorded by the WFP with those obtained from a gold-standard FP. While an exact correspondence was not anticipated, and good correlation, though not excellent, was expected, our hypothesis was that the findings would demonstrate the potential of this technology for portable gait analysis, providing an accessible and practical solution.

## Materials and methods

### ShokacShoes

The ShokacShoes are a newly developed sensorized shoe designed by Nakai et al. ^29^ and based on the ShokacChip ^30^. The ShokacChip is a small and thin 6-axis force-torque MEMS sensor. The sensor is able to measure vertical and shear forces as well as torque. Datasheet information is included in Table 1. Three ShokacChips were inserted in a standard shoe: one under the heel and two under the forefoot, as shown in Fig 1A and Fig 2A. The sensors were connected with a WR-FFC 0.50mm Type 1 FFC jumper cable and a WR-FPC SMT ZIF Horizontal Back Locking connector (Würth Elektronik eiSos GmbH & Co. KG, Deutschland) to the communication module, which included a daisy-chain Communication System and a bluetooth connection. The ShokacShoes also include an LSM6DS3 inertial measurement unit (STMicroelectronics, Geneva, Switzerland). All electronics were included inside an insole of 9 mm, making the ShokacShoes a potential WFP, highly comfortable and usable (Fig 2A).

**Fig 1.**
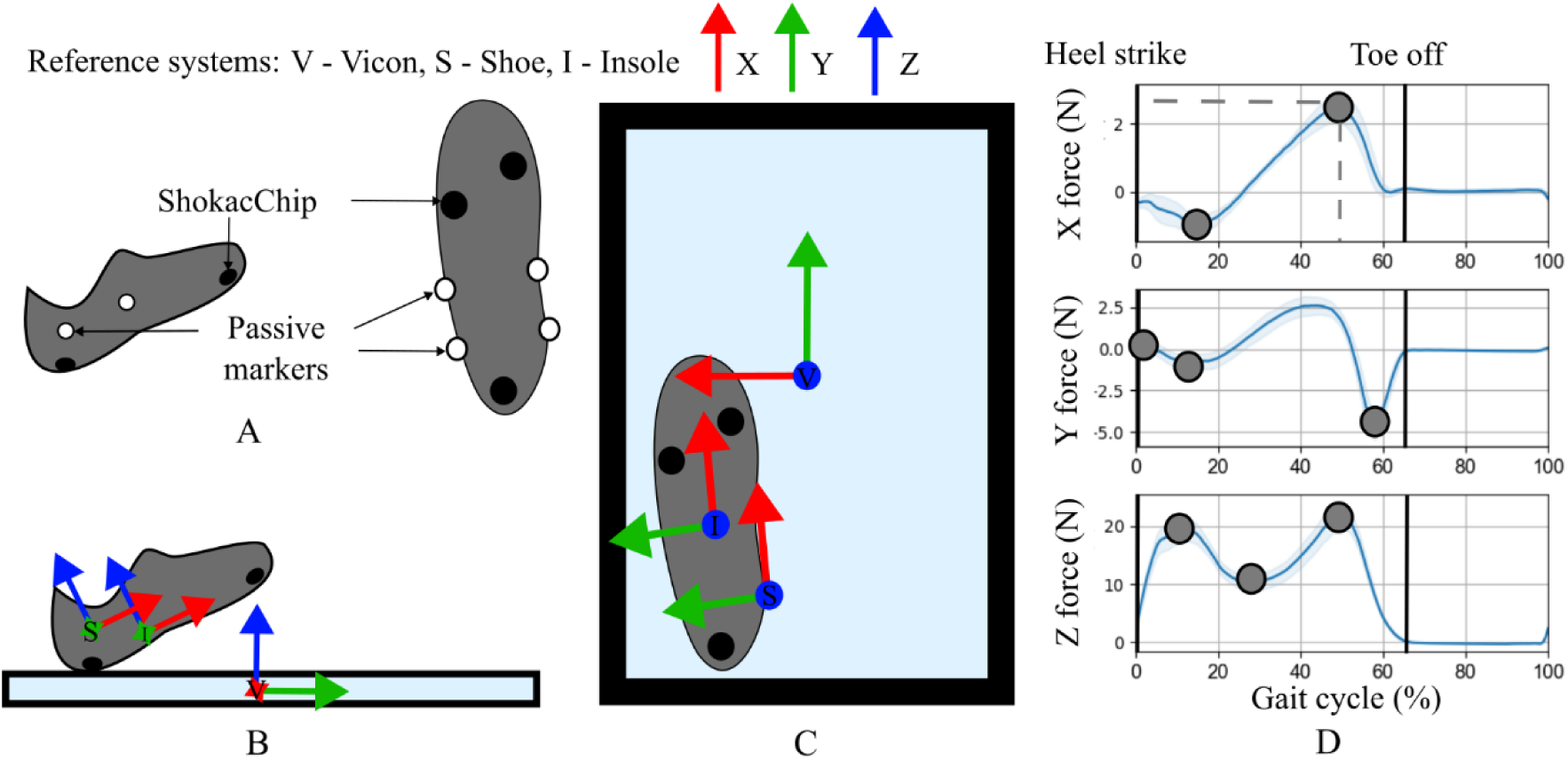
Graphical representation of the methodology process used to align the Vicon and ShokacShoes reference systems. A set of 4 passive markers were placed on the shoe to create a Shoe reference system (S). B. Lateral view of the Vicon (V), Shoe (S) and Insole (I) reference systems. C. Upper view of the reference systems. D. Force metrics calculated to compare WFP and FP (grey circles). Force metrics include the amplitude and the index as a % of Gait cycle. Heel strike and Toe off are used for computing the temporal parameters (Stride Times, Cadence and Stance Phase).

**Fig 2.**
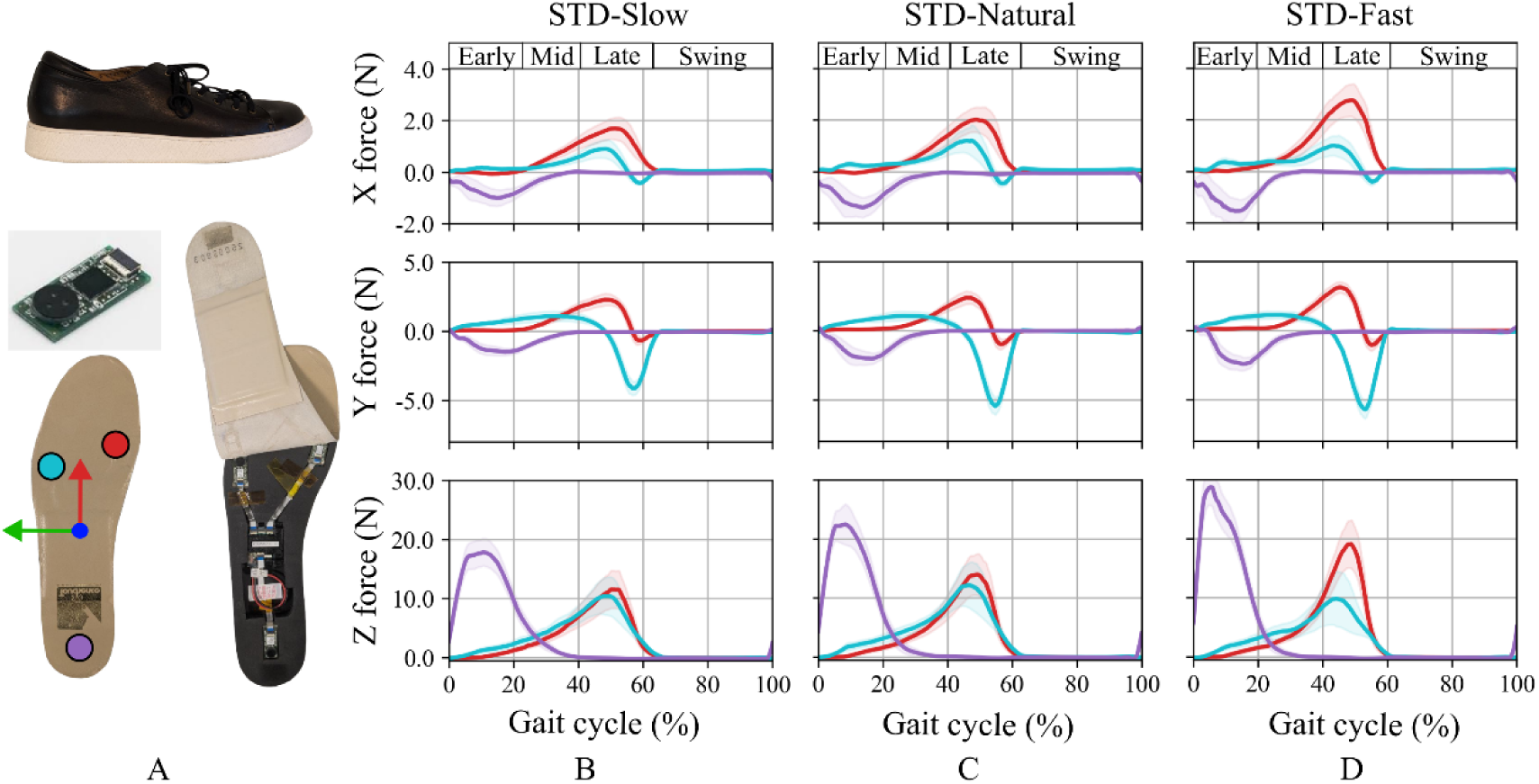
A. Final placement of the ShokacChip sensors inside a standard shoe with a 9-mm thick insole. Sensors are placed under the heel (purple sensor), the 1st metatarsal joint (red sensor) and the 5th metatarsal joint (blue sensor). The insole reference system is shown at the center of the insole (X in red, Y in green, Z in blue) B. Aggregated results of the forces recorded by each sensor during the slow walking condition with the standard insole (STD-Slow). The data from the 3 sensors were used to perform a threshold-based gait segmentation into Early, Mid, Late Stance, and Swing phases. C. Same as B for STD-Natural. D. Same as B for STD-Fast.

**Table 1.**
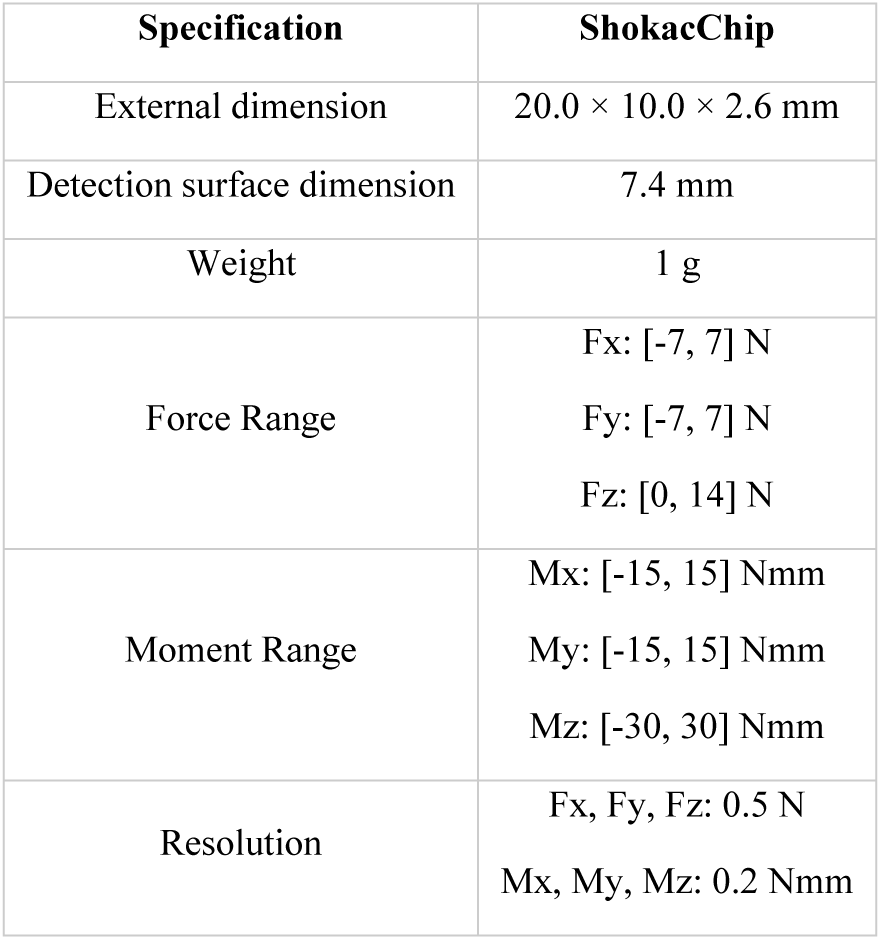
Technical specifications of the ShokacChip.

The data were recorded at 50 Hz by a custom program developed in Python 3.11. Recorded data included: three-dimensional forces and torque of the three ShokacChip sensors, three-dimensiona acceleration and angular velocity from the inertial measurement unit, the total three-dimensional forces of the shoe (calculated as the sum of the forces from the three sensors) and the center of pressure calculated based on the fixed position of the sensors in the shoe insole.

### Recruitment

Eight young healthy participants were enrolled in this study after signing the informed consent. Participants were included if they had a shoe size between 39 and 40 and did not present any traumatological, neurological or cardiological pathology that would compromise their walking abilities. Data were recorded in the frame of the project “Validation of the HYBRID platform for modeling and training gait”, accepted by the Ethics committee of Universidad Politécnica de Madrid and carried out in the Biomechanics Lab of the Universidad Politécnica de Madrid.

### Validation protocol

Participants were asked to walk in a 10-meter corridor wearing the ShokacShoes. The corridor was instrumented with 4 Vicon M2 mcam cameras operated by the Workstation 5.2.9 software (Vicon Motion Systems Ltd., Yarnton, UK) and two 9281E Kistler 60×40 cm force platforms (Kistler Group, Winterthur, Switzerland). Passive markers were placed on the lower segment of the participants following the Plug-In-Gait Vicon marker protocol.

Prior to data collection, participants completed a familiarization trial to warm up and identify an appropriate starting position that enabled natural foot placement on the force platforms without compensatory movements. Familiarization ended when participants felt comfortable, with a maximum of three trials per participant.

First, participants were asked to lift the shoes to perform the calibration and find the zero value of the WFP sensors. Then, participants were asked to stand on the border of the force platforms and kick a force platform with the left foot. This was used for data synchronisation (more details in section Data analysis). Next, participants walked until the beginning of the corridor, stopped for some seconds, and walked through the corridor. Finally, they walked back to the initial position out of the recording volume of the Vicon system and repeated the process 5 times. After walking 5 times, participants were asked to kick the force platform with the left foot again, as at the beginning of the trial. The 5 walking trials were repeated in 6 different conditions: three speed conditions and two insole conditions. Participants were asked to walk with an uncontrolled self-selected Slow, Natural, and Fast pace with the WFP with its standard insole (STD) and with an additional sneaker insole on top of it (SNK), to verify whether additional insoles (such as orthopedic insoles) could significantly modify the force measurements and whether these changes could have clinical implications. Speed instructions were provided as follows: “Walk at your natural, comfortable pace”; “Walk at a naturally slower pace, without exaggerating the slowdown so that your gait pattern remains natural”; and “Walk at a naturally faster pace, as if in a hurry, without running or attempting to walk as fast as possible”. The six walking conditions were performed in a fixed order: STD-Natural, STD-Slow, STD-Fast, SNK-Natural, SNK-Slow, and SNK-Fast, without randomization. This order was selected to avoid bias in the natural-speed condition and to allow Slow and Fast speeds to be defined relative to each participant’s natural gait, ensuring clear differences between conditions.

### Data analysis

Data were analysed in Python 3.12.6. Data from the WFP and the FP were manually synchronised based on the peak forces recorded by the FP (sampling frequency of 1080 Hz) and by the WFP (sampling frequency of 50 Hz). The synchronisation kicks were recorded at the beginning and at the end of the trial in order to accurately synchronise the systems in case of data loss. The data were synchronized based on the time difference between the peaks of the vertical force recorded by the left WFP and the FP.

The data from the right WFP were not analysed due to robustness issues. However, as the technology is identical, the results of the left side are expected to be generalizable to both shoes. The data from the FP, saved in the Vicon global reference system of Workstation (reference system *V* in Fig 1) were roto-translated to the reference system of the shoe (reference system *S*), which was created based on 4 passive markers placed on the shoe (Fig 1A). Then, the data were roto-translated again to the insole reference system (reference system *I*), the main reference system of the forces of the WFP. The *I* reference system was established at the center of the insole, rigid enough to ensure that the reference system was stationary. According to the *I* reference system, the main reference system from now on, the X direction is antero-posterior (anterior being +), the Y direction is medio-lateral (external being +), and the Z direction is vertical (upwards being +). This methodology allowed for a more direct comparison between FP and the WFP in identical reference systems. This reference system roto-translation was performed by automatically updating the model files inside the Workstation 5.2.9 program.

Then, gait events (heel strike and toe off) were retrieved separately for WFP and FP. For WFP, we implemented a semi-automatic threshold-based method. The threshold was subject-specific and condition-specific, and ranged from 0 to 2 N. The rising edge of the threshold crossing was used for detecting the heel strike and the falling edge to detect toe off. For the FP, heel strikes were manually retrieved based on the vertical GRF and the heel marker position. Toe offs were semi-automatically retrieved when the vertical GRF crossed a 5-N threshold.

Force and CoP data were segmented step by step based on the gait events, separately for the WFP and the FP. All steps were then interpolated to 1000 samples. Force data were compared between WFP and the FP by normalizing the force data from each sensor and each axis to the maximum force value of the median force of each subject. Normalization was performed to overcome the expected amplitude difference, taking into account the specifications of the sensors (maximum force range of 14 N). Then, the mean normalized forces were aggregated and meaned between subjects. Forces were statistically compared considering (1) 8 force metrics: the maximum and minimum value of X representing braking during heel strike and forward propulsion during toe off, the maximum value of Y representing medial loading during mid-stance, the 2 negative force peaks of Y representing lateral unloading during initial contact and pre-swing, the 2 peaks of Z representing vertical loading during weight acceptance and toe off and the force valley between the 2 peaks in the Z direction; (2) the position, as percentage of Gait Cycle, of each of the 8 force metrics; (3) gait temporal parameters such as step time in s, stance phase in % and cadence in strides/min for each system separately. Metrics are represented visually in Fig 1D.

### Statistical analysis

Statistical analysis were performed in Python 3.12.6. Differences between self-selected walking speeds were assessed using a repeated-measures ANOVA, followed by paired t-tests for post hoc comparisons with Holm–Bonferroni corrections. The effects of speed and insole using the WFP were quantified with the Wilcoxon signed-rank test, as the distribution was not normal for the majority of the metrics according to the Shapiro-Wilk test for normality. The Wilcoxon test was computed on paired samples to assess differences across conditions using the *stats* submodule of the *scipy* Python library with Holm-Bonferroni corrections. Similarly, effect size between conditions was quantified using Hedges’ g ^31^. The Wilcoxon signed-rank test, widely used in the state of the art, enables the comparison of our results with existing literature. However, given the repeated-measures design, the dependency of observations within subjects, and the hierarchical structure of the data, we complemented the Wilcoxon analysis with a linear mixed-effects model, which increases statistical power by accounting for random effects, and modeling variance components. The model included all main effects and their interactions and was specified as Value ∼ Speed ∗ Sensor with a random intercept for Subject to capture between-subject variability. A different model was built for each metric using the Python *statsmodels* module with a MixedLM implementation and with a Restricted Maximum Likelihood method.

To compare the normalized forces measured by FP and WFP, we used a Wilcoxon signed-rank test with Holm-Bonferroni corrections. The Wilcoxon test was run on the metrics obtained from the normalized forces for the force amplitudes and on the metrics obtained from the non-normalized forces for the temporal variables (indexes and gait parameters). Moreover, force data from FP and WFP were also temporarily compared by means of Statistical Parametric Mapping (SPM) using the *ttest2* function of the statistics module of the *spm1d* Python package. Moreover, linear regressions were used to study the relationship between non-normalized forces, indexes and temporal parameters measured by the FP and by WFP. Linear regressions were calculated with the *LinearRegression* class of the *sklearn* Python module from the data distribution of the non-normalized metrics. Intraclass correlation coefficients (ICC) were calculated using a two-way random-effects model with absolute agreement (ICC(2,1)), using the *intraclass_corr* function of the *pingouin* library. Agreement was evaluated using Bland-Altman analysis by calculating the mean difference (bias) and limits of agreement (mean difference± 1.96 × standard deviation of the differences) between WFP and FP. In order to increase the sample size, linear regressions, ICC and Bland-Altman analysis were calculated considering all the steps of all the subjects. This decision increased the sample size from 8 values (one per subject) to 40 (each subject performed 5 walking trials in each condition). Results were interpreted considering state-of-the-art ranges. R^2^ was considered as good to excellent from 0.76 to 1, as moderate to good from 0.51 to 0.75, as fair from 0.26 to 0.50 and as none to little below 0.25 ^32^; ICC was considered as moderate from 0.50 to 0.75, as good from 0.75 to 0.90 and as excellent over 0.90 ^33^.

## Results

### Recruitment and data recording

Eight subjects with a shoe size between 39 and 40 were recruited for this study (7 females, 1 male). Mean age was 22.1 ± 3.5, mean weight was 57.5 ± 4.3 kg and mean height was 165.6 ± 2.4 cm. Walking speed was 0.94 ±0.19 m/s during Slow walking, 1.39 ± 0.18 m/s during Natural walking and 1.77 ± 0.13 m/s during Fast walking. A significant effect of speed was observed (F = 97.7, p < 0.001). Post hoc comparisons revealed significant differences between natural and fast walking (p < 0.001) and between natural and slow walking (p < 0.001). Due to technical issues with the Vicon system, data were lost during the STD-Fast condition of Subject 01 and STD-Natural condition of Subject 02. For these conditions, the aggregated data include the data of only 7 subjects. For the other 4 conditions, data are aggregated between the 8 subjects recruited for the study.

### Effects of speed, insole condition and number of steps selected

Fig 2 shows the ShokacChip sensor placement inside the ShokacShoes (Fig 2A) and the corresponding force profiles aggregated between subjects and recorded under three walking conditions: STD-Slow, STD-Natural, and STD-Fast (Figure 2B, C and D respectively). During early stance (0 - 20 % of gait cycle), GRF are primarily concentrated under the heel (purple sensor). The X, Y and Z forces under the heel increased by 61, 65 and 63 % respectively when increasing walking speed from STD-Slow to STD-Fast. During mid stance (20 - 40 % of gait cycle), load distribution is uniform across the three sensors. During late stance (40 - 60 % of gait cycle), weight is mainly supported by the sensors under the 1st (red sensor) and 5th metatarsal joint (blue sensor). X force, primarily observed under the 1st metatarsal joint, increased by 61 % from STD-Slow to STD-Fast. Y force, mainly observed under the 5th metatarsal joint (purple sensor), increased by 32 % from STD-Slow to STD-Fast.

Fig 3 shows the resulting forces aggregated between subjects considering only the total forces (the sum of the X, Y, and Z forces measured by the 3 sensors of Fig 2). Data are shown as a function of speed (Fig 3A), and as a function of the insole (Fig 3B). Figure 3C shows the boxplots of the distribution of 9 relevant metrics: the values and indexes of maximum of the X force, the minimum of the Y force, the 1st maximum of the Z force; and temporal parameters Stride Time, Cadence and Stance Phase. Results show that subjects walked with a natural cadence of 52.5%, a stance phase of 61.7 % and a maximum vertical load of 19.6 N. When walking faster (STD-Fast), in median, subjects reduced their stance phase by 3.4 % (p-value 0.06, Hedges’ g −1.7), increased their cadence by 15.8 % (p-value 0.06, Hedges’ g 2.6) and increased their maximum vertical load by 31% (p-value 0.3, Hedges’ g 0.44) compared to the STD-Natural condition. When walking slower (STD-Slow), in median, subjects increased their stance phase by 3.3 % (p-value 0.2, Hedges’ g 0.7), reduced their cadence by 20.6 % (p-value 0.06, Hedges’ g −1.6), and reduced the maximum vertical load by 9.2 % (p-value 0.06, Hedges’ g 1.4) compared to the STD-Natural.

**Fig 3.**
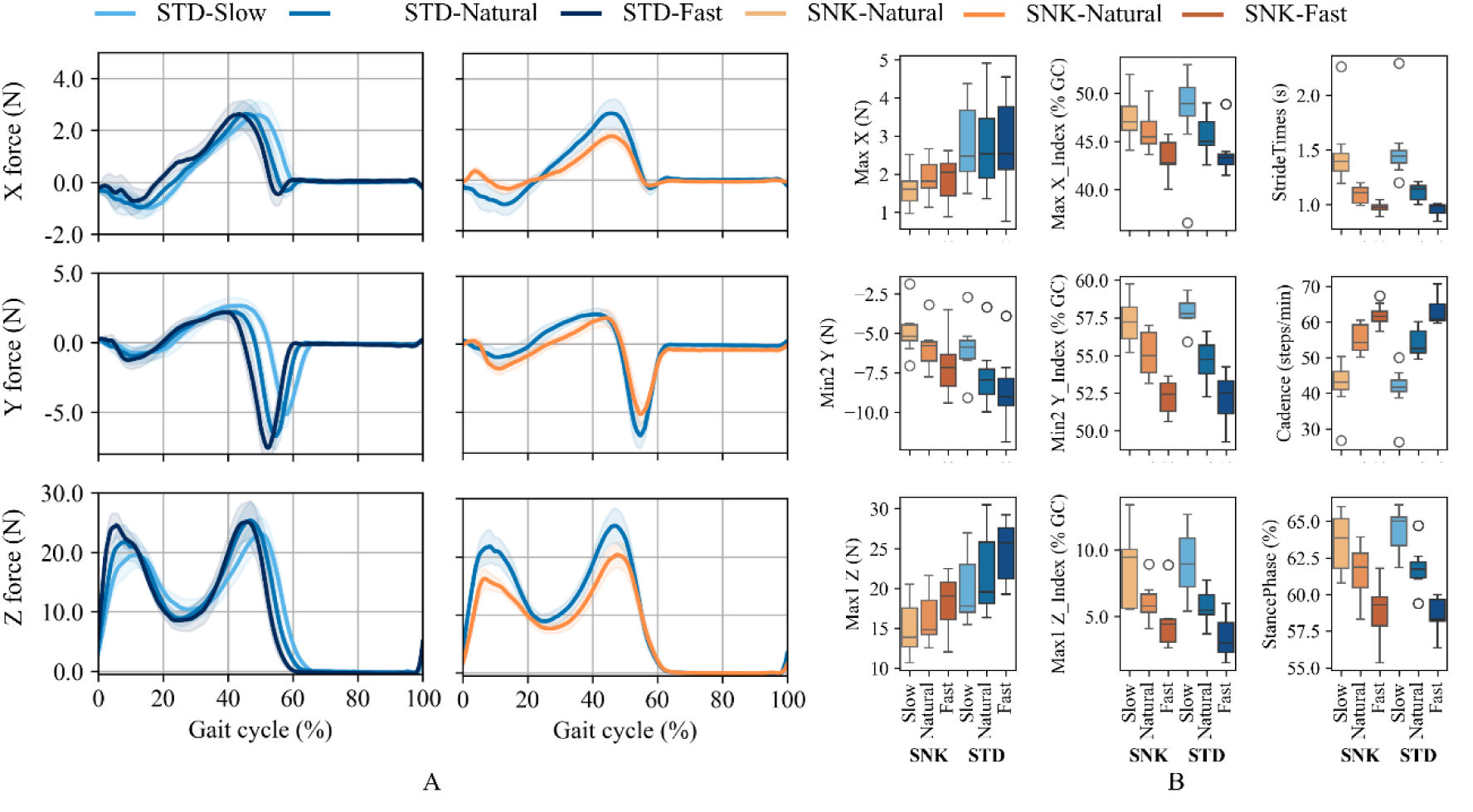
A. Aggregated results of the forces measured by the WFP. On the left side, the figure shows the X, Y and Z forces of the STD condition when walking at Slow, Natural and Fast speeds. On the right side, the figure shows the forces of the STD condition compared to the SNK condition at a natural speed. B. Boxplots of the distribution of 9 relevant metrics throughout the 6 conditions: SNK-Slow, SNK-Natural, SNK-Fast, STD-Slow, STD-Natural, and STD-Fast.

When using the additional sneaker insole (SNK-Natural) on top of the sensors, maximum amplitude was significantly reduced by 28.4 % (p-value 0.06, Hedges’ g −0.9), 37.3 % (p-value 0.05, Hedges’ g 1.13) and 24.2 % (p-value 0.03, Hedges’ g −1.8) respectively for X, Y and Z directions compared to the standard condition (STD-Natural). However, the use of the SNK insole did not have a significant impact on indexes or spatiotemporal variables. The numerical values of the most relevant metrics are included in Table 2.

**Table 2.**
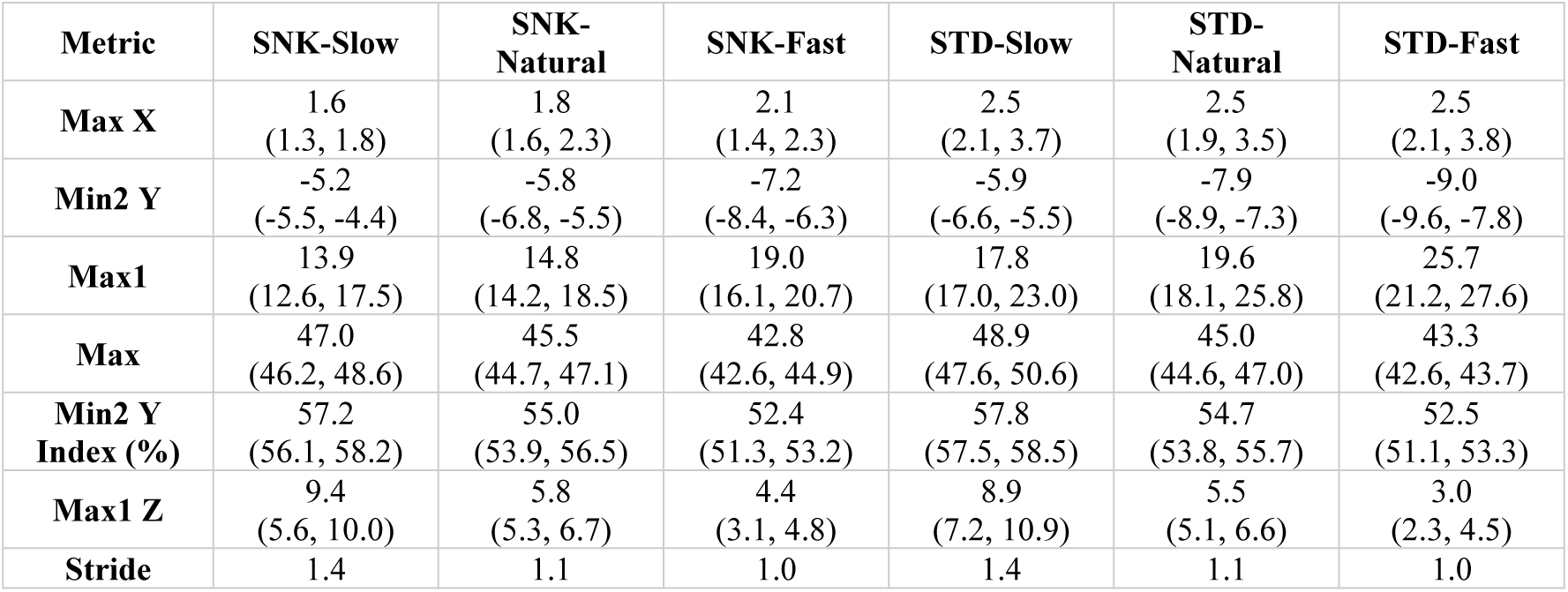
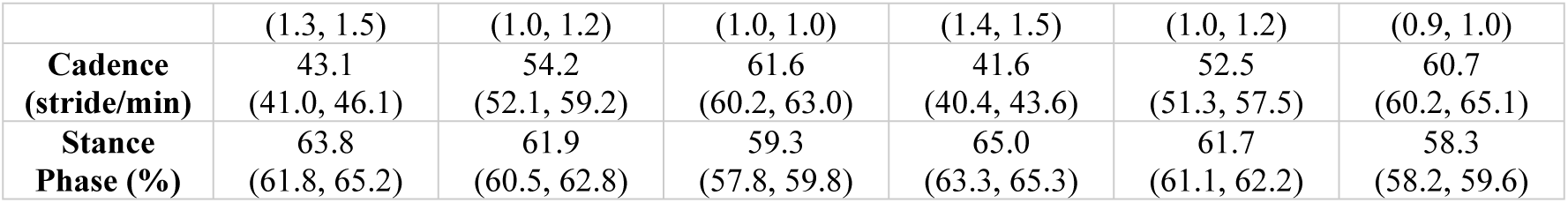
Summary of computed gait metrics for the six walking conditions using the data measured by the WFP. Metrics are shown as Median (Q1-Q3).

Similarly to the Wilcoxon test, the LMM showed in Fig 4 demonstrates that changing the speed (from Natural to Slow or from Natural to Fast) has a significant effect on all temporal variables. The row Sp:Slow and Sp:Fast of Fig 4 shows statistically significant differences on the indexes of X, Y and Z, and on Stride Times, Cadence and Stance Phase. Changing the speed also modified the amplitude of the Y and Z forces, but not of the X force. Regarding the effect of the insole, the row Sh:SNK shows that the SNK insole has a significant effect on the amplitude of the forces (in all axes X, Y and Z) but not on temporal metrics. The numerical results of all calculated metrics, as well as the p-values of the Wilcoxon signed-rank test are included as Supplementary materials.

**Fig 4.**
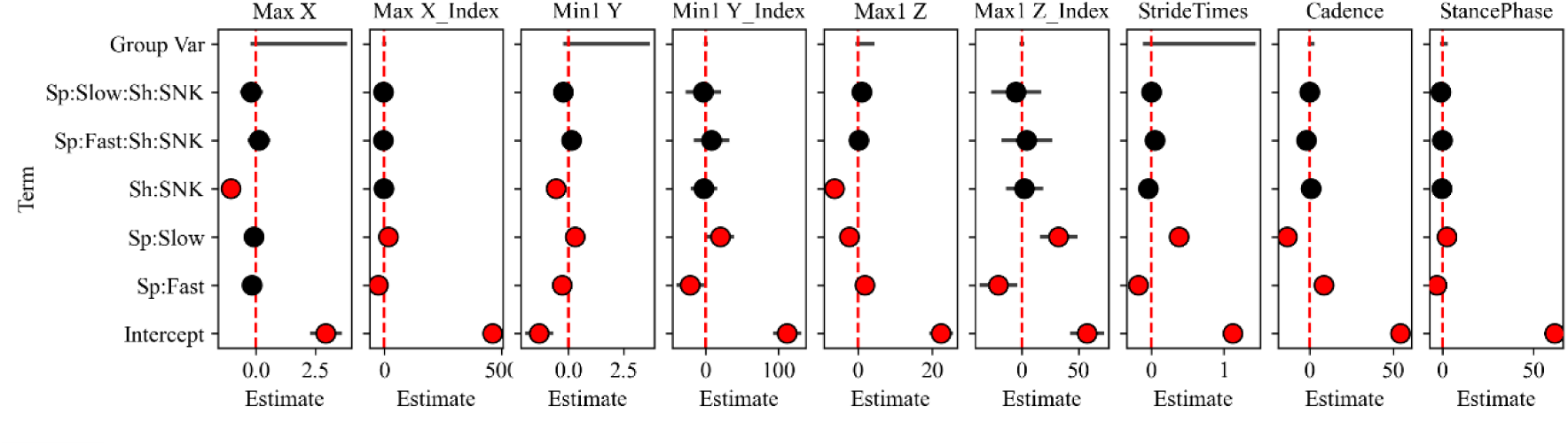
Results of the Linear Mixed Model. The labels on the y axis represent the effects tested with the model. Sp stands for Speed, Sh for Shoe. The effects of the insole type (SNK or STD) and of speed (Slow, Natural or Fast) were tested by comparing the different conditions against the “standard” condition, STD-Natural. Effects with statistically-significant p-values under 0.05 are shown in red.

### Validation against gold-standard

Fig 5 shows the comparison between the data measured by WFP and by the FP, considering the measured three-dimensional GRF (Fig 5A) and its normalized version (Fig 5B) in the *I* reference system for the aggregated STD-Natural condition, considered as a representative condition. Considering the aggregated data between subjects, the forces measured by the FP ranged from −91.3 to 168.1 N in X direction, −214.3 to 68.4 N in Y direction and 0 to 663.8 N in Z direction. Regarding the WFP, forces ranged from −1.1 to 2.5 N in X direction, −7.9 to 0.7 N in Y direction and 0 to 19.6 N in Z direction. Figure 5C shows the boxplot distribution of 9 relevant metrics measured with FP and WFP.

**Fig 5.**
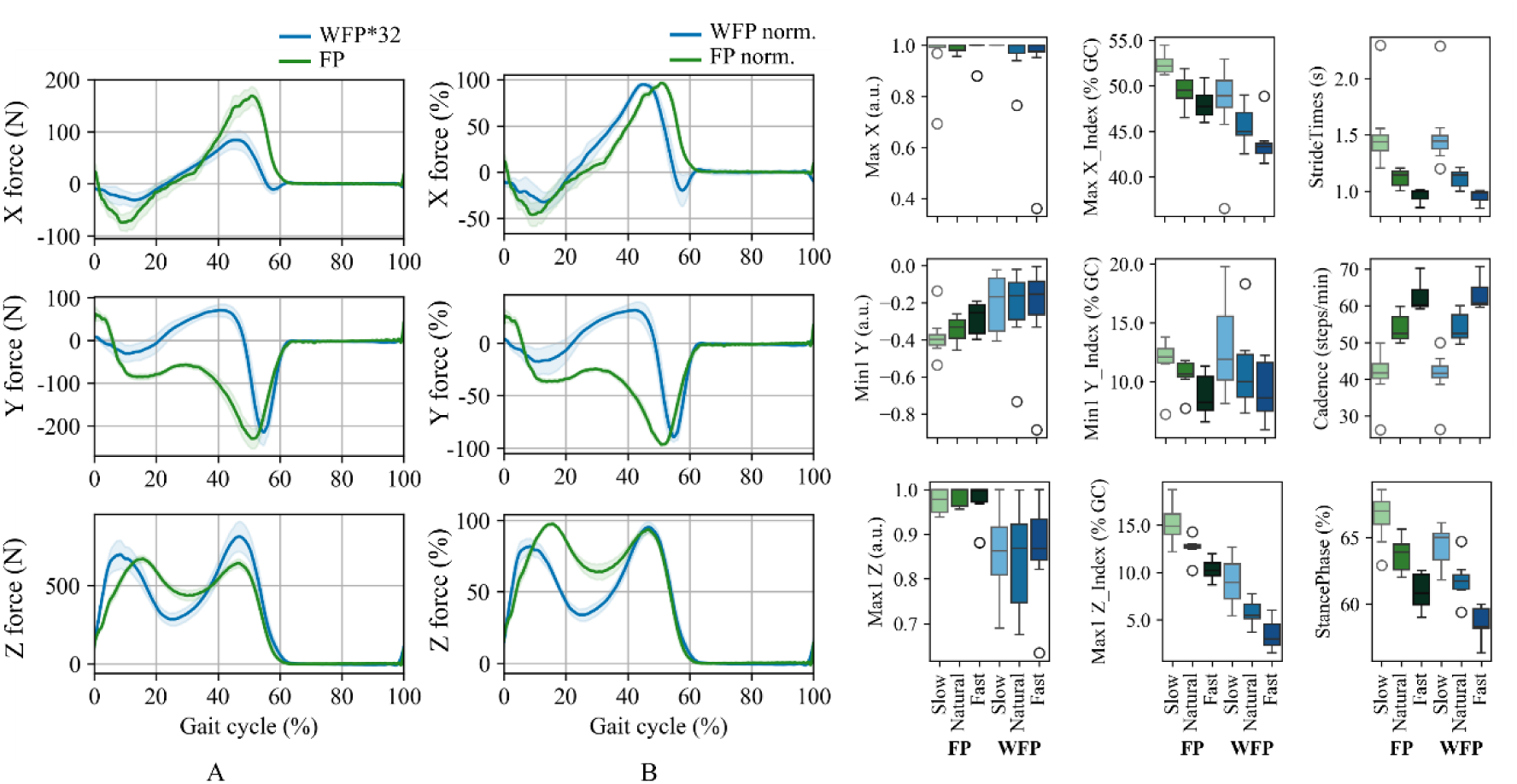
Comparison between the WFP (blue) and the FP (green). A. Three-dimensional forces of the STD-Natural condition of the non-normalized forces. WFP data are multiplied by 32. B. Three-dimensional forces of the STD-Natural condition of the normalized forces. C. Boxplots of the distribution of 9 relevant metrics throughout three conditions: STD-Slow, STD-Natural, STD-Fast.

Significant changes were observed only in the maximum values of the Y and Z forces, even after normalization. Considering the STD-Natural as a representative condition, the Max1 Z value was of 1.0 when measured by the FP and 0.87 when measured by the WFP (p-value 0.05, Hedges’ g −1.0). The vertical force Z includes two peaks, the first at around 12.5 % of gait cycle and the second at around 50% of gait cycle. The normalization was carried out with the maximum Z force, which corresponded to Max1 Z for FP and Max2 Z for WFP. This difference in the normalization value explains the 0.13 difference in the Max1 Z score. A similar result was obtained for the Y force: significant changes were observed between WFP and FP (p-value 0.03, Hedges’ g −2.6). Regarding temporal variables, some significant differences were also observed in the indices of the metrics. The difference between FP and WFP was of 0.3%of gait cycle (p-value 0.3, Hedges’ g −1.2), 1.8%(p-value 0.03, Hedges’ g 3.3) and 6 % of gait cycle (p-value 0.03, Hedges’ g −3.6) respectively for Max X, Min2 Y and Max1 Z direction. However, no significant differences were observed at the level of Stride Times (p-value 0.77, Hedges’ g −0.3) or Cadence (p-value 0.92, Hedges’ g 0.3). All numerical values of the most relevant metrics are included in Table 3.

**Table 3.** Summary of computed gait metrics for STD conditions at different walking speeds using the data measured by the FP and the WFP. Metrics are shown as Median (Q1-Q3).

| Metric | FP Slow | FP Natural | FP Fast | WFP Slow | WFP Natural | WFP Fast |
| --- | --- | --- | --- | --- | --- | --- |
| **Max** | 1.0<br>(0.97, 1.0) | 1.0<br>(0.98, 1.0) | 1.0<br>(1.0, 1.0) | 1.0<br>(0.99, 1.0) | 1.0<br>(0.98, 1.0) | 1.0<br>(1.0, 1.0) |
| **Min** | -0.37<br>(-0.42, -0.31) | -0.38<br>(-0.42, -0.36) | -0.33<br>(-0.38, -0.30) | -0.40<br>(-0.43, -0.37) | -0.33<br>(-0.39, -0.29) | -0.25<br>(-0.37, -0.21) |
| **Max1 Z** | 0.97<br>(0.96, 0.98) | 1.0<br>(0.98, 1.0) | 1.0<br>(0.98, 1.0) | 0.98<br>(0.95, 1.0) | 1.0<br>(0.96, 1.0) | 1.0<br>(0.97, 1.0) |
| **Max X Index** | 52.2<br>(51.6, 52.9) | 49.5<br>(48.6, 50.6) | 47.7<br>(46.8, 49.0) | 48.9<br>(47.6, 50.6) | 45.0<br>(44.6, 47.0) | 43.3<br>(42.6, 43.7) |
| **Min1 Y Index (%)** | 12.1<br>(11.6, 12.8) | 10.7<br>(10.4, 11.6) | 8.2<br>(7.6, 10.5) | 11.9<br>(10.1, 15.5) | 10.0<br>(8.7, 12.3) | 8.6<br>(7.5, 11.6) |
| **Min2 Y Index (%)** | 55.0<br>(53.9, 55.5) | 51.1<br>(50.6, 52.0) | 49.3<br>(48.1, 50.2) | 57.8<br>(57.5, 58.5) | 54.7<br>(53.8, 55.7) | 52.5<br>(51.1, 53.3) |
| **Max1 Z** | 14.9<br>(14.0, 16.2) | 12.8<br>(12.6, 12.9) | 10.2<br>(9.6, 11.0) | 8.9<br>(7.2, 10.9) | 5.5<br>(5.1, 6.6) | 3.0<br>(2.3, 4.5) |
| **Stride** | 1.4<br>(1.4, 1.5) | 1.1<br>(1.1, 1.2) | 1.0<br>(0.9, 1.0) | 1.4<br>(1.4, 1.5) | 1.1<br>(1.0, 1.2) | 1.0<br>(0.9, 1.0) |
| **Cadence**<br>**(strides/min)** | 41.8<br>(40.2, 44.2) | 52.4<br>(51.1, 57.0) | 60.3<br>(59.9, 64.4) | 41.6<br>(40.4, 43.6) | 52.5<br>(51.3, 57.5) | 60.7<br>(60.2, 65.1) |
| **Stance Phase**<br>**(%)** | 67.0<br>(66.0, 67.7) | 63.9<br>(62.6, 64.5) | 60.8<br>(60.0, 62.2) | 65.0<br>(63.3, 65.3) | 61.7<br>(61.1, 62.2) | 58.3<br>(58.2, 59.6) |

Furthermore, FP and WFP were also compared at a temporal level with an SPM analysis (Fig 6A). SPM shows that significant differences occurred between 34.5 and 44.4 % of gait cycle for X forces, between 0.6 and 3.7 %, between 22.4 and 50.6 %, between 93.4 and 94.2 % and between 96.4 and 96.9 % of gait cycle for Y force, and between 12.4 and 29.4 % of gait cycle for Z force. Regarding the linear regressions between FP and WFP (Fig 6B), R^2^ values showed excellent correlation and ICC values showed moderate agreement for Stride Times and Cadence. For Stride Times, R^2^ is equal to 0.97, RMSE to 0.013 and ICC to 0.65. For Cadence, R^2^ is equal to 0.97, RMSE to 0.63 and ICC to 0.66. Good agreement was observed for Max Y (R^2^ 0.61, ICC 0.77), Min2 Y Index (R^2^ 0.52, ICC 0.78) and Max1 Z Index (R^2^ 0.04, ICC 0.81) and excellent for Valley Z (R^2^ 0.00, ICC 0.92). For other metrics, no correlations and agreement were observed (R^2^ < 0.25 or ICC < 0.5). Bland-Altman plots show a slight proportional bias for X, Y and Z amplitudes and indexes. Stride times and Cadence show minimal bias and a strong linear agreement. However, stance phase is less precise, with a negative bias of around 2%.

**Fig 6.**
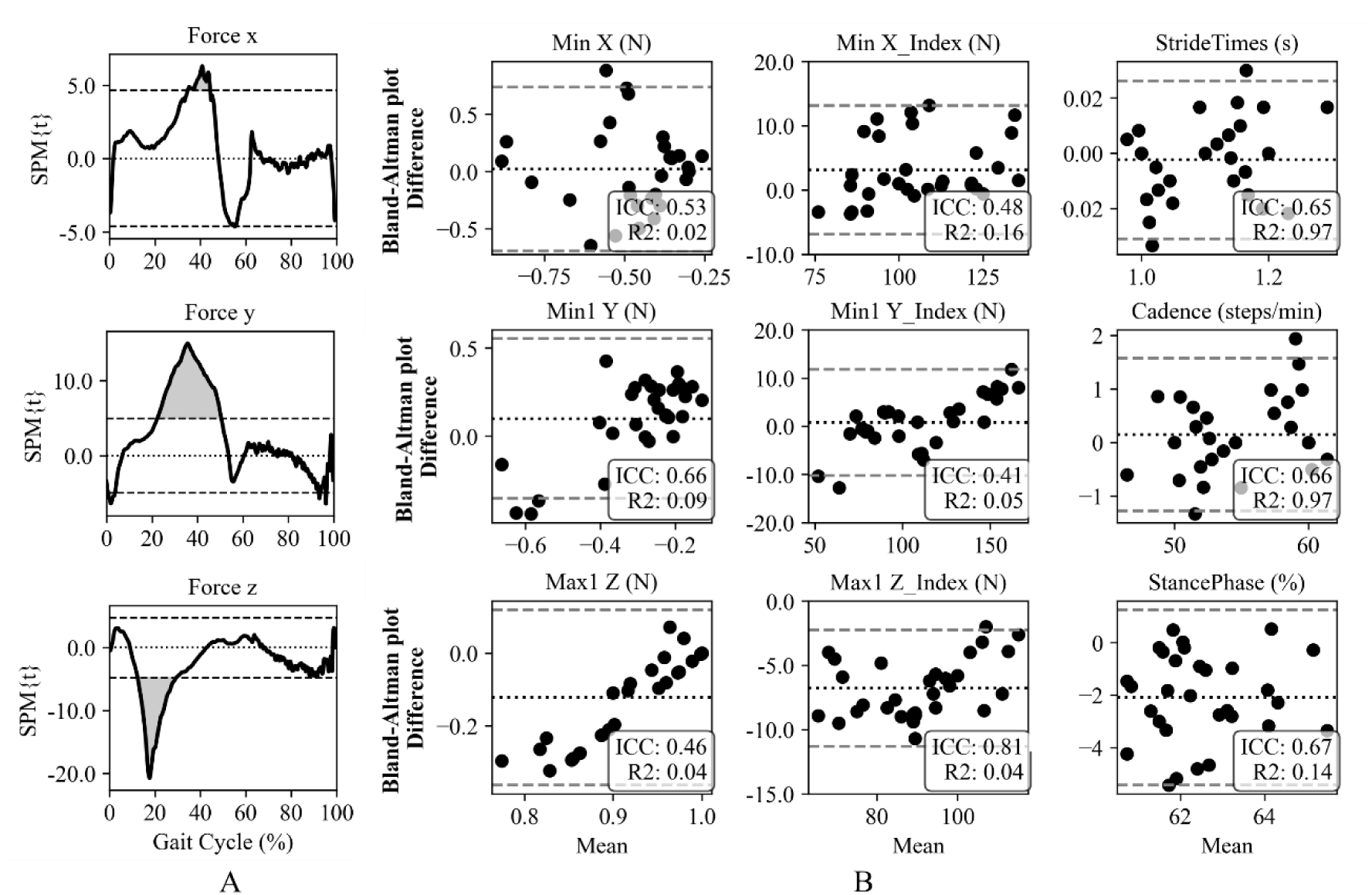
Statistical analysis comparing the forces measured by the FP and the WFP. A. Statistical Parametric Mapping. B. Bland-Altman plots including the results of the ICC and linear regression (R^2^).

The numerical results of all calculated metrics, as well as the uncorrected and corrected pvalues of the Wilcoxon signed-rank test, the Hedges’g effect size, the *R*^2^ values of the linear regressions and the ICC values are included as Supplementary materials.

## Discussion

This work presents a feasibility study proving that gait analysis can be carried out with the Shokac-Shoes, a novel WFP designed to overcome the inherent limitations of conventional FP. Traditional FP confine gait assessments to controlled laboratory settings and restrict the number of analyzable steps to the quantity of embedded FP within a walkway. Current state-of-the-art solutions for overcoming these limitations are sensorized footwear which primarily comprises (1) sensorized insoles which offer high usability but only estimate vertical forces rather than capturing three-dimensional force components (OpenGo, F-Sacl Go, Medilogic…) and (2) sensorized shoes, capable of measuring three-dimensional forces but which rely on bulky structures or wired connections, thereby reducing practicality ^12,24,27^. To the best of the authors’ knowledge, the ShokacShoes are the first solution in the field to integrate a thin, sensorized insole that can measure three-dimensional force components within any commercial shoe while employing a wireless communication protocol. This configuration enables seamless data transmission and provides a highly usable, unobtrusive device for gait biomechanics analysis beyond laboratory constraints, even in home settings.

In our study we presented a feasibility study of a biomechanical gait analysis with the ShokacShoes. This WFP integrates three sensors located under the heel, the 1st metatarsal joint, and the 5th metatarsal joint, enabling accurate segmentation of the gait cycle into Early, Mid, and Late stance phases, as well as the swing phase. Our findings indicate that this segmentation is highly precise as subphase timing varied consistently with changes in walking speed. Such granularity is less direct with traditional FP which rely on a single sensing area. Sensor placement is also critical for providing relevant biomechanical information. Incorporating two sensors in the forefoot region allows for a more precise understanding of the forefoot load distribution, which is essential for identifying pathological gait patterns ^34^. Some studies have previously reported pressure distribution, not three-dimensional forces, under different segments of the foot using sensorized insoles ^35^. Moreover, the inherent design of the state-of-the-art sensorized shoes able to measure three-dimensional forces, which only integrate one sensor under the forefoot, does not allow to study forefoot load distribution. This new WFP could thus have a more extended diagnostic potential than the state of the art. We envision its application across the continuum of care from prodromal stages of Alzheimer’s and Parkinson’s disease to home-based rehabilitation following traumatic or neurological injuries.

Moreover in this study, we demonstrated that walking speed and insole type significantly influence respectively temporal parameters and force amplitudes. As anticipated, increasing speed primarily affected the position of force indices, stride duration, cadence, and the percentage of stance phase, which aligns well with previous biomechanical findings ^36^. When an additional SNK insole was placed over the sensors, force amplitudes were significantly reduced. This outcome was expected, as the foam composition of the SNK insole redistributed the forces applied to the sensors, thus reducing the sensor reading. Nevertheless, this modification did not significantly alter temporal parameters. The purpose of adding the SNK insole was to assess whether individuals using orthopedic insoles or requiring soft, protective insoles (such as those at risk of pressure ulcers) could still benefit from the WFP. Although the general recommendation is to use the standard configuration of the WFP, with direct contact with the sensors, results indicate that the WFP remains functional even with supplementary insoles supporting its usability in diverse clinical and everyday-life scenarios. The performance of the WFP was evaluated against the gold-standard Kistler FP in a preliminary biomechanical analysis. We compared force amplitude and waveform characteristics, force-related indices, and temporal gait parameters. We did not obtain an exact match across all metrics. Linear regression analyses revealed excellent R^2^ only for stride time and cadence, and good or excellent agreements were observed in a limited number of metrics, throughout temporal parameters, forces amplitudes and indexes positions. Only good agreement was observed for stride time and cadence. Additionally, the limited measurement range of the WFP sensors prevented a direct force comparison, requiring normalization procedures that reduced the interpretability of the results. Still after normalization, force amplitude showed statistically significant differences between FP and WFP.

Statistically significant differences were also observed in the X- and Z-direction indices. Overall, the results indicate moderate to good agreement for temporal metrics, whereas the accuracy of ground reaction force measurements, assessed against the ground truth provided by static force plates, remains limited. These discrepancies may be attributed to sensor positioning: the heel sensor is located approximately 2.5 cm from the posterior end of the shoe while the sensor beneath the 1st metatarsal joint lies about 7.5 cm from the anterior tip. Consequently, during the very early stance phase and the terminal portion of late stance, the WFP may fail to capture forces accurately which might have a clinical impact. Clinically meaningful variations in stance phase are around 2 to 4 %. For example, differences of approximately 2.2% have been reported between admission and discharge in stroke survivors ^37^, and differences of around 4 % occur when individuals with Parkinson’s disease discontinue levodopa medication ^38^. The clinical implications of the numerical differences of the forces and gait temporal parameters between FP and WFP should be analysed in the future in more specific biomechanical studies. Nevertheless, these differences did not affect higher-level parameters such as cadence or stride time. Furthermore, accurate biomechanical interpretation of GRF depends not only on quantitative metrics but also on the quantitative assessment of signal morphology. With the SPM analysis, we observed that some significant differences occur mainly in Y and Z directions but only during midstance. These differences may be attributed to the limited number of sensors embedded in the shoe. While integrating three sensors, one under the heel and two under the forefoot, which enables the assessment of forefoot load distribution, the midfoot lacks coverage, substantially reducing the forces recorded by the WFP compared to the FP during midstance. Ideally, full-sole sensor coverage would be required to avoid underestimation of forces ^39^. Future design iterations could incorporate an additional sensor on the lateral aspect of the foot. Although this modification would introduce additional technical complexity, such as additional wiring to the Bluetooth module, the potential improvement in accuracy would justify the effort.

Our preliminary biomechanical comparison between FP and WFP aligns with previous validation studies of state-of-the-art sensorized shoes, which typically benchmark shoe outputs against goldstandard FP measurements ^12,24,27,40,41^. However, protocols for these comparisons vary widely across studies. Some works validate raw force measurements using Root Mean Squared Error, considering the similar force ranges between sensorized shoes and FP ^12,24,27,28^. In contrast, given the inherent amplitude differences in our data, we opted for a more robust approach: we statistically compared peak forces through a Wilcoxon signed-rank test and curve morphology through an SPM analysis, as seen in other studies ^42^. The combination of the Wilcoxon test and the SPM analysis not only provides an error metric to quantify force measurement accuracy but also captures temporal differences between systems. To complement this, we also derived gait temporal parameters which are clinically relevant metrics that are rarely addressed in such validation studies. Moreover, some studies have explored center of pressure and balance metrics ^21,41^ or estimated joint moments via inverse dynamics ^28,43^. Specifically, Snyder et al. ^28^ embedded the same ShokacChip sensors in a different shoe and although in their study they did not compare the data from the sensorized shoe to a gold-standard FP, they used the measured GRF to estimate knee adduction moments with the two systems. Given that our study represents a preliminary biomechanical analysis, we focused our comparison on force-derived metrics.

However, further development should include a more comprehensive set of metrics. Our study has several limitations: (1) The recording protocol included only eight participants, which adequate for a feasibility assessment. However, this limited sample size prevents the results from being generalized to a broader population. It also does not eliminate the need for a proper validation study with a statistically determined sample size. (2) Data were recorded from only one shoe due to technical limitations. These issues should be resolved prior to the validation study to enable the calculation of additional metrics, such as temporal symmetry and double support time. (3) Walking speed was uncontrolled across participants and no randomization was performed.

Although all subjects were healthy young adults with relatively narrow walking speed variability, and a minimal risk of fatigue after completing 30 ten-meter corridors (six conditions with five trials each), these limitations should be addressed in future studies to increase the statistical power of the results; (4) the low force amplitude inherent to the sensor design combined with their number, placement, and the 50 Hz sampling frequency, prevented a straight-forward comparison with the raw data of the gold standard FP. Enhancing the ability of the ShokacShoes to function as a WFP might require addressing these constraints with an iterative design process; (5) in this study, we analyzed only the steps performed on the platforms rather than examining all steps along the corridor which is a capability offered by the ShokacShoes, thanks to their portability, that is not available in traditional FP. Including additional steps in future analyses could provide more reliable insights into gait variability in individuals with neurological disorders. As these limitations reduce the overall strength of this feasibility study, we will target future research by involving larger cohorts, additional metrics, full-corridor analysis, and including neurologically-impaired participants to explore the clinical applicability of the shoes in a subsequent validation study. Despite these constraints, we believe this work is relevant to the scientific community because (1) ShokacShoes are the first sensorized shoes capable of measuring three-dimensional GRF wirelessly in a usable shoe, (2) the force curve shape and derived temporal parameters align well with gold-standard FP, (3) the shoes incorporate more sensors than FP, enhancing diagnostic granularity and (4) the ShokacShoes overcome FP limitations, enabling their use in less controlled environments. Expanding this study with new data will help demonstrate the potential of ShokacShoes as a wearable force platform that could replace traditional FP in daily biomechanical gait analysis and in uncontrolled scenarios.

## Conclusion

This study demonstrates the feasibility of using the ShokacShoes as a WFP for measuring threedimensional GRF. The prototype incorporates a thin insole with three sensors and wireless data transmission and showed a reliable performance across different speeds and conditions with expected speed- and insole-dependent force patterns. While comparisons with conventional FP were encouraging, discrepancies highlight the need of an iterative design refinement to promote measurement accuracy. Future work will focus on validating the applicability of a new version of the ShokacShoes against gold-standard FP in a comprehensive study involving diverse real-world scenarios and pathological conditions. These efforts will be critical to establishing ShokacShoes as a robust tool for replacing traditional FP and assessing biomechanics even outside laboratory settings.

## Data Availability

All numerical results are available as Supplementary Material.

## Acknowledgements

We would like to thank all the participants of the study.

## Supporting information

Supplementary_material.xlsx. Document including additional numerical results.

